# How Low Could Semaglutide Prices Fall? An Analysis of Production Cost and Implications for Global Access Ahead of Patent Expiry

**DOI:** 10.64898/2026.03.04.26347508

**Authors:** Jacob Levi, Samuel Cross, Nydile Ramesh, Francois Venter, Andrew Hill

## Abstract

**Objectives:** To estimate potential launch prices of generic semaglutide following patent expiry from 2026 and to quantify the global obesity and type 2 diabetes (T2DM) burden in countries where generic access may become possible.

**Methods:** We used World Bank population data and World Obesity and Diabetes Atlas prevalence estimates to calculate obesity and T2DM burden in countries where semaglutide patents expire in 2026 or were not filed. Patent status was identified using MedsPaL and cross-checked with regional databases. We updated established cost-plus pricing methodologies using 2024-2025 Indian API shipment data to estimate production costs for oral and injectable semaglutide, incorporating formulation, packaging, taxation, and profit assumptions.

**Results:** Ten countries with 2026 patent expiry represent 44% of the global population and 48% of the global obesity burden. No patent filings were identified in 150 additional countries. By the end of 2026, generic injectable semaglutide could be distributed in 160 countries where 69% of global T2DM and 84% of clinical obesity occurs. Estimated generic injectable costs ranged from $28–140 per person-year, while oral formulations ranged from $186–380 per person-year. Injection devices contributed disproportionately to total cost.

**Conclusion:** Patent expiry could substantially expand access to semaglutide at dramatically lower prices, particularly in high-burden settings. However, device costs, secondary patents, and health system constraints may limit equitable uptake without coordinated policy action.

**Study Importance:** *What is already known about this subject?:*

- Semaglutide is highly effective for obesity and cardiometabolic disease but remains unaffordable in many low- and middle-income countries due to high branded prices and patent protections.
- Previous cost-plus analyses show that generic competition can substantially reduce prices of essential medicines after patent expiry.

*What are the new findings in your manuscript?:*

- Using 2024–2025 API shipment data, we estimate generic injectable semaglutide could be produced for $28–140 per person-year following 2026 patent expiry.
- By 2026, generic semaglutide could be available in 160 countries comprising 69% of global T2DM and 84% of clinical obesity burden.

*How might your results change the direction of research or the focus of clinical practice?:*

- Provides an evidence base for procurement planning and price negotiations ahead of patent expiry.
- Highlights the importance of addressing device costs and secondary patents to ensure equitable global access.

## Introduction

Overweight and obesity are a growing global public health challenge, with 2.5 billion adults affected worldwide (1). Although 1 in 8 people globally live with obesity (BMI >30kg/m^2^), around 70% of the burden falls on low- and middle-income countries (LMICs) (2). In 2022, WHO estimated that above-optimal BMI contributed to 3.7 million deaths from noncommunicable diseases (NCDs), including cardiovascular disease, type 2 diabetes (T2DM), cancers, and chronic respiratory disease (1). NCDs now account for 82% of premature deaths (before age 70) in LMICs, compounding their ongoing infectious disease burden (2), effects both influenced by the globalisation of neoliberal market frameworks which entrench health disparities through its influence on trade, policy, and the social and commercial determinants of health (3). In this context, expanding access to effective weight-management therapies - including GLP-1 receptor agonists such as semaglutide - has important clinical and public health implications.

Semaglutide, initially developed for glycaemic control in T2DM, has demonstrated substantial metabolic, cardiovascular, and renal benefits in people both with and without diabetes (4,5). Now recognised as an effective obesity therapy and included in the WHO Essential Medicines List, it lowers HbA1c and improves glycaemic control for people with T2DM (6), whilst also reducing weight, waist circumference, blood pressure, lipids, and inflammatory markers (7,8). In obesity without diabetes, weekly semaglutide 2.4 mg produces sustained mean weight loss of approximately 15% over 104 weeks versus 2.6% with placebo (7). Trials such as SUSTAIN-6 and FLOW show significant reductions in nephropathy progression and major kidney disease events, alongside reductions in major adverse cardiovascular events, improved heart failure outcomes, and lower all-cause mortality in high-risk populations (6,9).

However, it is important to highlight that GLP-1 agonists are not a panacea for obesity. A recent meta-analysis of 37 studies including over 9,000 patients found that although weight loss of up to 20% can be achieved during treatment, weight regain after discontinuation is rapid (averaging 0.8 kg per month) and may be faster than with lifestyle-based interventions or some oral therapies (10). As a result, individuals stopping GLP-1 agonists, such as semaglutide, may return to their pre-treatment weight within approximately 18 months, exacerbating the chronic and relapsing nature of obesity. These findings raise important questions about long-term treatment duration, adherence, affordability, and expectation management, particularly in low-resource settings. Further research is needed to evaluate the recurrence of cardiometabolic risk following weight regain. Semaglutide and other anti-obesity medications should therefore be considered components of a sustained, comprehensive weight-management strategy rather than short-term or standalone solutions.

Semaglutide was granted FDA approval for treatment of T2DM in 2017 (as Ozempic) and obesity in 2021 (as Wegovy). However, despite its clinical promise, the high price of branded semaglutide -$6,936-7,716 per person-year (ppy) for oral semaglutide and $4,248-9,648ppy for injectable formulations in the USA, prior to recent price increases of 3.5-3.5% in 2025 (11) - has significantly limited its access in LMICs (12), and uptake in high-income countries has been slow due to reimbursement restrictions and out-of-pocket costs to users (13). Profits from Novo Nordisk’s semaglutide drugs (Ozempic and Wegovy) and of tirzepatide, the GLP-1 agonist produced by its main competitor Eli Lilly, are staggering. In 2024, Novo Nordisk made over $26 billion in revenue from Wegovy and Ozempic, whilst tirzepatide (brand names Mounjaro and Zepbound) had combined sales of nearly $16 billion. Ozempic and Wegovy ranked as the third and sixteenth highest-grossing pharmaceuticals worldwide in 2024 respectively, as measured by total sales revenue (14).

Patents for semaglutide are due to expire in several countries from April 2026 – examples include India, China, Canada, Brazil and Turkey - enabling the development and distribution of generic formulations. This shift in intellectual property rights could significantly reduce semaglutide prices and expand access globally, through generic production and pooled procurement, mirroring earlier successes with generic antivirals (15) and other essential medicines (16). Such changes are particularly important if LMICs are to benefit from the clinical benefits semaglutide confers.

## Methods

We used World Bank population estimates and morbidity data from World Obesity and Diabetes Atlas to estimate the number of people with obesity and T2DM in countries where generic semaglutide may become available. We used the Medicines Patent Pool database (MedsPaL) to determine where semaglutide patents are held and when they are due to expire and cross checked with regional databases where necessary.

We updated previously established and published cost plus (cost+) pricing methodologies (16) to estimate the cost of production for semaglutide. We reviewed The Trade Vision LLC database, examining records from November 2024 to November 2025 of semaglutide active pharmaceutical ingredient (API) imports and exports to and from India, providing shipment-level data on quantity, price, and origin. Thirty-three shipments of API were extracted, and a weighted mean cost/kg was calculated. Production costs were estimated using established cost+ methodologies, incorporating API costs and estimates for formulation, packaging, labour-costs, transportation, taxation on profit (of 27%), and profit margins (of 30%), assuming efficient large-scale generic manufacture. A full list of cost assumptions with references can be found in supplementary table 1. The weighted mean cost/kg of API was multiplied by different approved dosing regimens for oral and injectable semaglutide.

## Results

Figure 1 shows the countries where semaglutide will become generically available in 2026. These 10 countries, shown in orange, carry 47% of the obesity burden worldwide and 49% of T2DM. No patent filings were identified in 150 other countries. By the end of 2026 injectable generic semaglutide could be distributed in 160 countries (orange and green countries), where 69% of T2DM and 84% of clinical obesity occurs.

**Figure 1.**
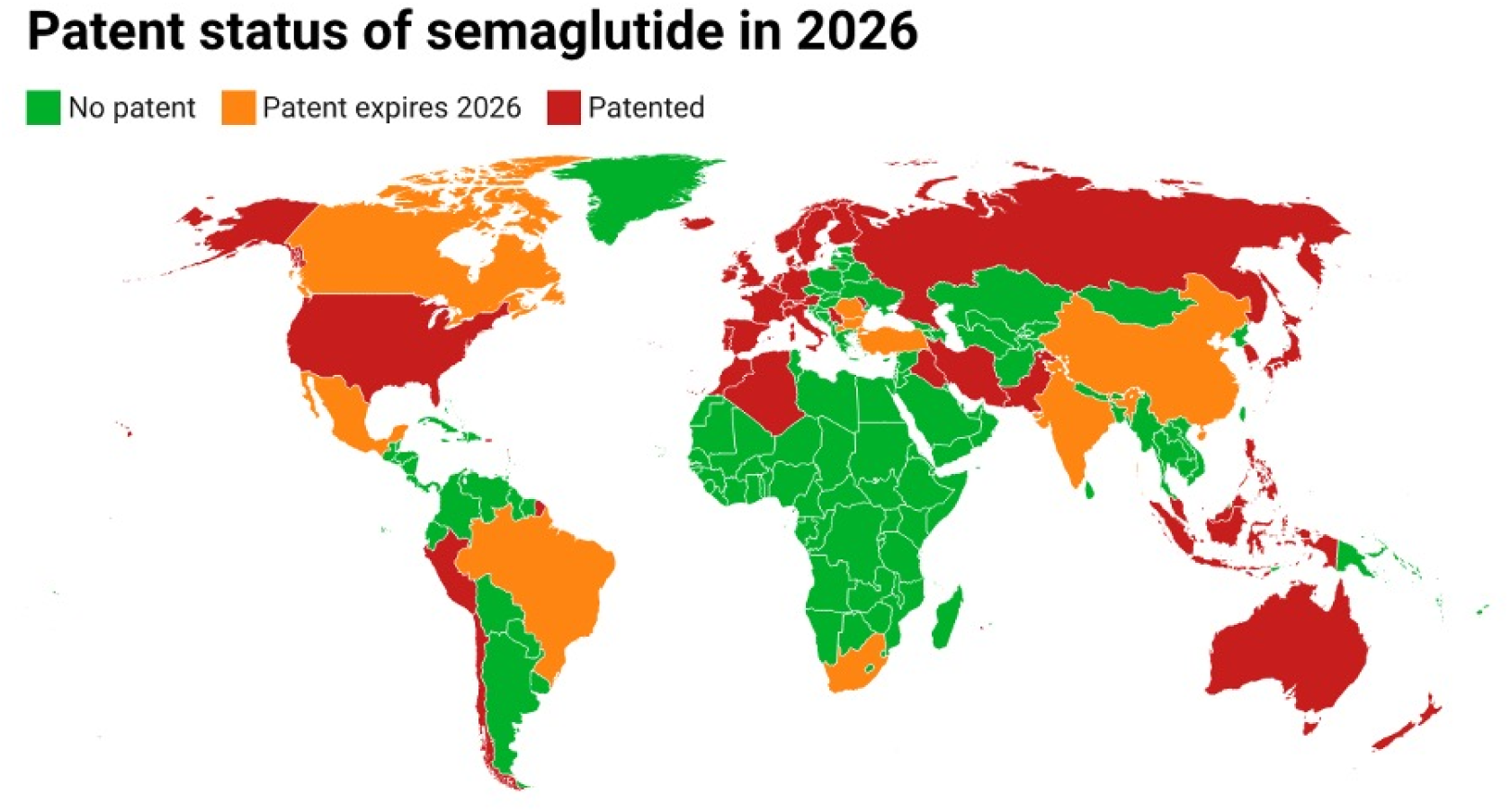
Choropleth map to show countries in which semaglutide patents expire in 2026. Using cost+ methodology, generic semaglutide could become available for just $28-134ppy for the establishment of a treatment naive patient on 1.7mg once per week treatment, or $29-138ppy for the establishment of a treatment naive patient on the higher dose of 2.4mg once per week (figure 2a). Once patients are fully established on treatment, injectable treatment is near identical, ranging from $28-135ppy for low dose treatment (1.7mg per week) and $30-140ppy for higher dose injectable treatment (2.4mg once per week).

**Figure 2a.**
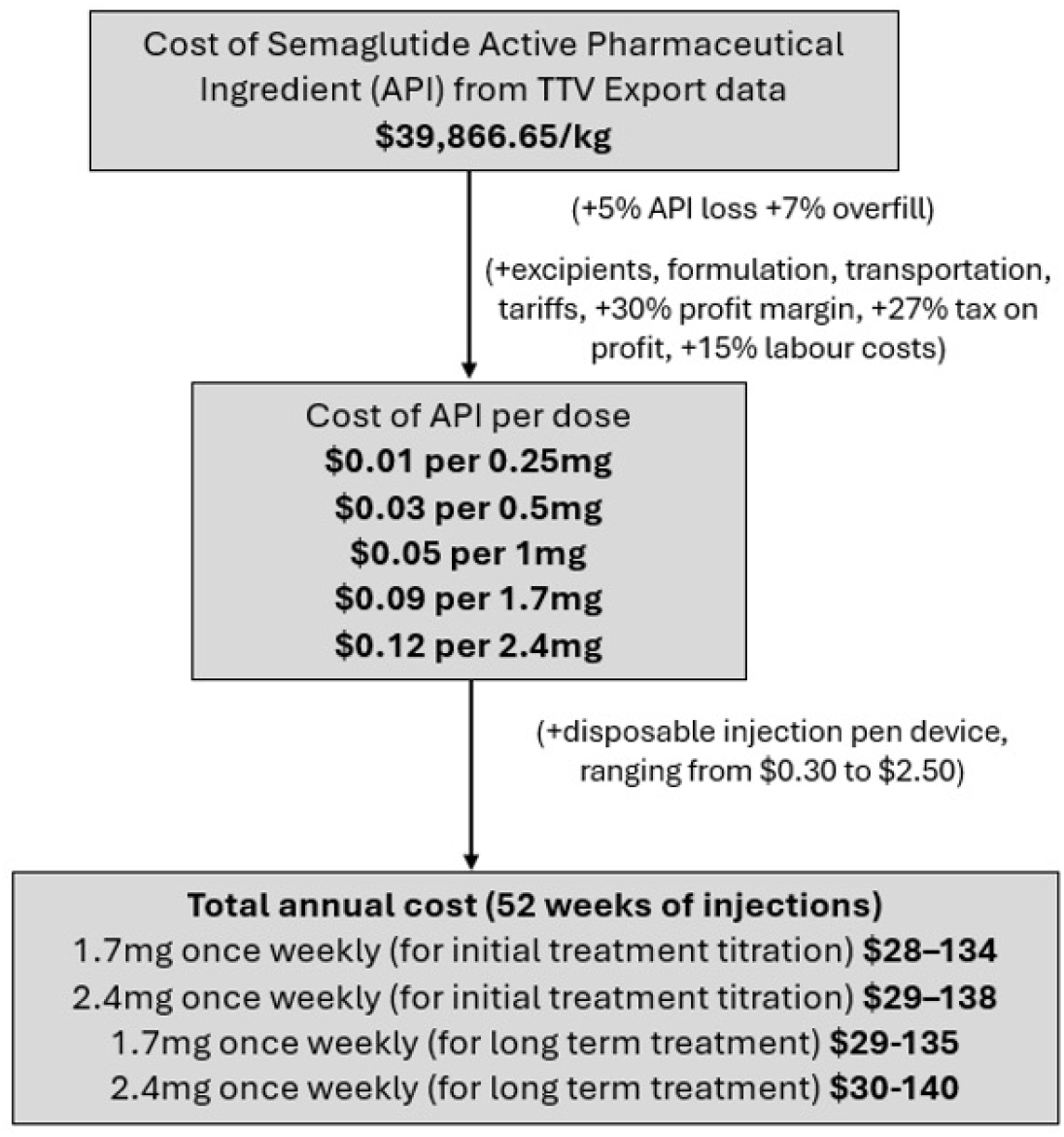
Cost+ analysis for injectable semaglutide treatment courses. Oral treatment courses are more expensive, as much higher doses of the API is needed per week (figure 2b). For treatment naive patients, generic oral semaglutide could become available for around $186ppy for those taking 7mg once per day, and $344ppy for those taking 14mg once per day. For patients that are already established on treatment, each subsequent year of oral therapy would cost $193ppy for 7mg once per day tablets, and $380ppy for high dose treatment of 14mg once per day.

**Figure 2b.**
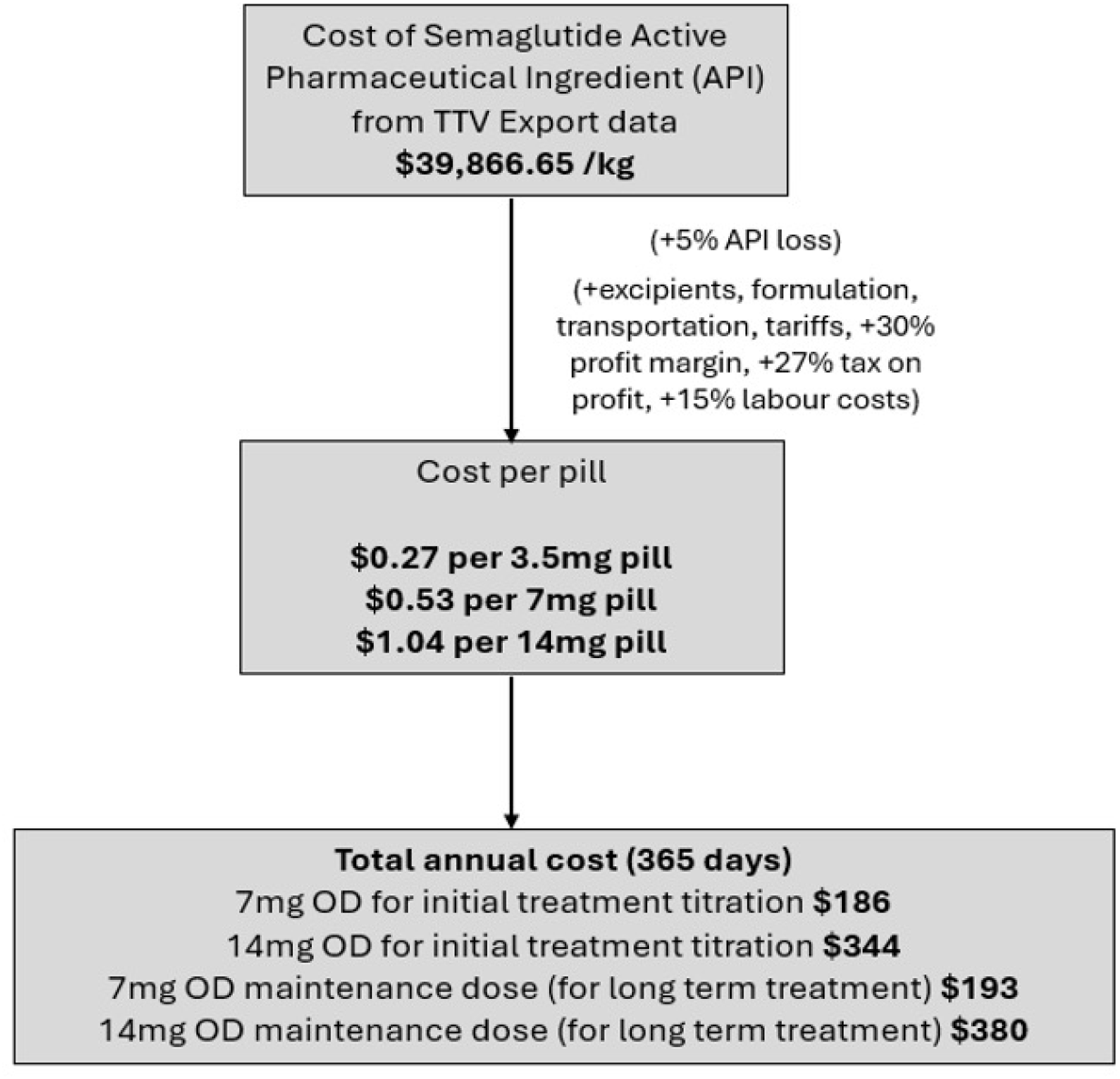
Cost+ for oral semaglutide treatment course. Production cost analysis indicates that API costs are low - around $0.01 (0.25mg) to $0.12 (2.4mg) per dose. In contrast, disposable injection pens cost $0.30-$2.50 per device (figure 3), meaning the device alone can cost 8-68 times more than the API plus all other production costs combined.

**Figure 3.**
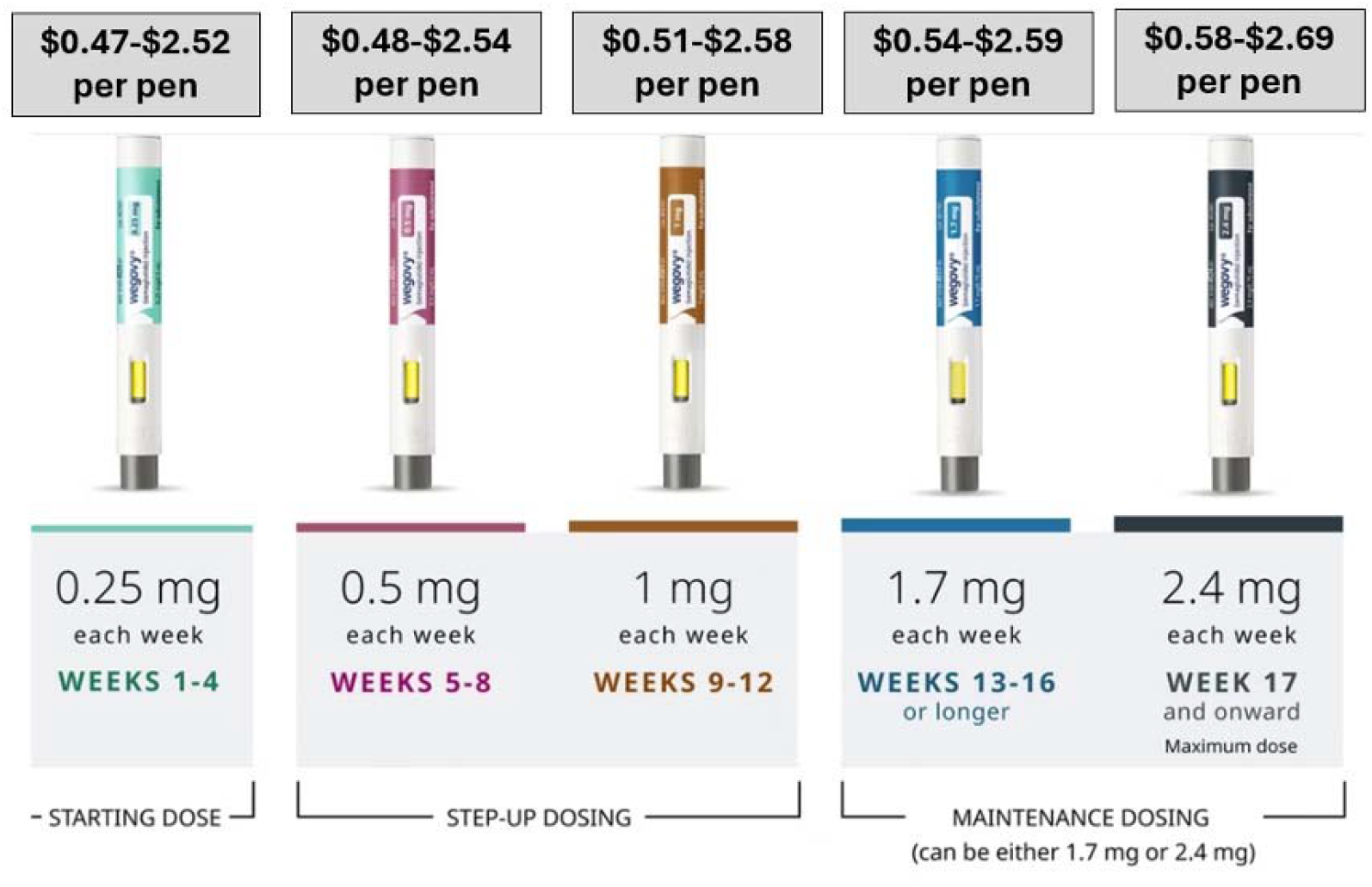
Cost per injectable semaglutide pen.

## Discussion

Several limitations should be acknowledged. First and foremost, although we used established cost-plus price assumptions, actual production costs will vary by manufacturer, scale, and procurement conditions. Secondly, shipment prices from India capture current market transactions, not necessarily the lowest achievable cost under mature, high-volume global generic production – API prices usually fall over time with introduction of generic competition resulting in lower production costs (17). Lastly, injection pens represent a major share of total cost, yet pricing data are less transparent than API data. Since 52 pens are needed per year (1 per week), the affordability of generic semaglutide will depend largely on low-cost device mass production. Even then, annual obesity treatment is likely to remain more expensive than first-line anti-retroviral drugs in some LMICs.

Although core semaglutide patents begin expiring in 2026, Novo Nordisk has constructed an extensive “patent thicket” of more than 20 patent families comprising 220 patents, across 28 countries, many focused on formulations and particularly delivery devices, with protections extending to 2033. Recent analysis suggests many GLP-1 patents focus on devices rather than the drug itself, raising concerns that patents may function primarily to extend market exclusivity (18). Intellectual property challenges from companies including Viatris and Huadong Medicine, which argued that semaglutide is too structurally similar to liraglutide to justify extended protection, have so far been unsuccessful (19). Meanwhile, generic momentum is building: Indian and Chinese manufacturers are advancing late-stage development, including oral formulations, and competition is expected to intensify after patent expiry (20).

Limited access to semaglutide risks widening global health inequalities. Without affordable pricing models or generic competition, these therapies will remain concentrated in high-income settings despite disproportionate need elsewhere. Lower prices through generic access could enable earlier intervention for obesity, integration into primary care, and a shift from treating advanced metabolic complications toward preventing long-term morbidity. However, generic entry alone is not a panacea. Budget stagnation, reduced external funding, procurement barriers, and health system constraints may limit uptake even if prices fall. Furthermore, long-term safety, durability of weight loss and associated cardiometabolic benefits, and real-world effectiveness in LMIC populations require continued evaluation and surveillance.

Importantly, pharmacotherapy cannot address the structural determinants driving obesity, including food insecurity, poverty, urbanisation, and commercial food environments. Sustainable impact will require coordinated policy approaches that combine equitable medicine pricing, strengthened primary care systems, lifestyle support, and broader public health interventions. Without deliberate action on affordability and implementation, the benefits of novel anti-obesity therapies are unlikely to reach the populations bearing the greatest burden.

## Conclusion

Using updated API shipment data and cost-plus pricing methods, we estimate that generic semaglutide could launch from 2026 at a fraction of current branded prices - with injectable formulations particularly affordable – in 160 countries comprising 69% of T2DM and 84% of clinical obesity globally. These projections are likely conservative as API costs typically fall further with scale and competition. However, device costs, secondary patents, financing constraints, and health system barriers may limit equitable access without coordinated policy and procurement action.

## Data Availability

All data produced are available online and upon reasonable request to the authors.

