## Supplementary table 1 for "How Low Could Semaglutide Prices Fall? An Analysis of Production Cost and Implications for Global Access Ahead of Patent Expiry"

**Supplementary Table 1 – References for fixed cost assumptions used in cost+ modelling**

| Tableting, formulation, coating | $0.01 | Hill AM, Barber MJ, Gotham D. Estimated costs of production and potential prices for the WHO Essential Medicines List. BMJ global health. 2018 Jan 1;3(1):e000571. |
| --- | --- | --- |
| Excipient costs | Formula 2 x API mass x 2.63 | Hill AM, Barber MJ, Gotham D. Estimated costs of production and potential prices for the WHO Essential Medicines List. BMJ global health. 2018 Jan 1;3(1):e000571. |
| Packaging per box | $0.10 | Barber MJ, Gotham D, Bygrave H, Cepuch C. Estimated Sustainable Cost-Based Prices for Diabetes Medicines. JAMA Network Open. 2024 Mar 4;7(3):e243474-. |
| Labour | 11% | Fairhead C, Fortunak J, Layne J, Johnson M, Smalley S, Lutterodt A, et al. 174. Generic Lenacapavir HIV Pre-Exposure Prophylaxis could be Produced for $25 Per Person Per Year. Open Forum Infect Dis 2026;13:ofaf695.004. https://doi.org/10.1093/ofid/ofaf695.004. |
| Transportation and tariffs | 15% | Hill A, Levi J, Fairhead C, Pilkington V, Wang J, Johnson M, et al. Lenacapavir to prevent HIV infection: current prices versus estimated costs of production. Journal of Antimicrobial Chemotherapy 2024;79:2906–15. https://doi.org/10.1093/jac/dkae305. |
| Tax | 27% | Standard Indian tax rates for generic suppliers  India - Corporate - Taxes on corporate income n.d. https://taxsummaries.pwc.com/india/corporate/taxes-on-corporate-income (accessed February 17, 2026). |
